# Mathematical Model to Study Early COVID-19 Transmission Dynamics in Sri Lanka

**DOI:** 10.1101/2020.04.27.20082537

**Authors:** Sanjeewa Nishantha Perera, Naleen C Ganegoda, Dhammika Deepani Siriwardhana, Manuj C Weerasinghe

**Affiliations:** Research & Development Centre for Mathematical Modeling, Department of Mathematics, University of Colombo, Colombo 03, Sri Lanka; Department of Mathematics, University of Sri Jayewardenepura, Nugegoda, Sri Lanka; Department of Disability Studies, Faculty of Medicine, University of Kelaniya, Ragama, Sri Lanka; Department of Community Medicine, Faculty of Medicine, University of Colombo, Colombo 08, Sri Lanka

## Abstract

**Background:** World Health Organization declared COVID-19 as a pandemic on 11^th^ March. Sri Lanka is currently experiencing a cluster epidemic with a specific group of overseas returnees and their contacts. The objective of this study was to develop a mathematical model to predict the epidemic in Sri Lanka incorporating measures taken for social distancing and prevention of social gatherings.

**Methods:** A hybrid model incorporating both exponential and polynomial features was developed and parameters were estimated. The developed model was validated using the datasets of three reference countries. Finally, the model was applied to the Sri Lankan data to simulate the epidemic behaviour. Additional features were incorporated to the model to examine the effects of current control measures.

**Findings:** Sri Lanka will have a peak of 177 COVID-19 active cases at the end of second incubation period from the index case of our projection, if the same trend continues. At 10% risk, we project a peak of 263 COVID-19 active cases at the end of third incubation period, and a peak of 353 at the end of fourth incubation period. Should the risk level reach 20%, the peak will be above 1000 active cases after 90 days. Simulations incorporating control measures predict that, deviation from the control measures currently in place could trigger exponential behaviour of the epidemic.

**Interpretation:** The hybrid model combining exponential and polynomial functions showed promising results to predict COVID-19 epidemic. Projections indicate that any early relaxation of control measures is not advisable. This methodological approach can be replicated in other settings at the initial stages of the epidemic.

## Introduction

The global COVID-19 caseload had exceeded over one million and sixty thousand deaths by 4^th^ of April 2020 [1]. It is still spreading across countries and communities. Health systems are overwhelmed due to the high caseload. Hence, lowering and flattening of the epidemic peak is particularly important, as it reduces the acute pressure on healthcare systems [2].

Several methods of community mitigation strategies are heavily advocated as the key prevention methods. These include cancellation and suspension of events with super spreader potential, use of social distancing measures to reduce direct and close contact between people in the community, travel restrictions including reduced flights, suspension of public transport and route restrictions without compromising the work of essential services, voluntary home quarantine of contacts, and clear communication (e.g. on aspects of hygiene) from health authorities to ensure verified information to the community [3].

Projections on the behaviour of an epidemic is extremely useful in planning the responses of the community, health services, and in general all sectors directly and indirectly partnering in the mitigation process. A few mathematical models and projections presented to date have attempted to understand the COVID-19 pandemic in detail, particularly based on data from cases in Wuhan in China (where the virus began) and from those returning from China to other countries. Those models have concentrated on early dynamics of transmission, epidemic behaviour with the application of different preventive measures, and infectivity dynamics of the pathogen [2,4,5]. In addition, attempts to predict resource need in healthcare to face the challenges of COVID-19 are emerging [6].

Sri Lanka is in a unique position in this epidemic; being an island with a population of 21 million. Sri Lanka provides free of charge healthcare at the point of delivery to the total population in the state hospitals. Currently, there are 628 state hospitals providing indoor healthcare facilities with the capacity of 83,275 beds. This makes 3.9 hospital beds per 1,000 population [7]. However, the majority of Sri Lankan hospitals provide non-specialised primary care and have very limited facilities and staff to manage conditions at an advanced stage such as severe respiratory distress. The preventive health care system in Sri Lanka is relatively strong with a dedicated field health staff, functioning under 347 Medical Officer of Health [MOH] areas which cover the entire country. Prevention, notification and control action on communicable diseases is one of the key functions of the MOHs. Hence, the ability to rapidly deploy health staff at the community level during an epidemic situation is a positive feature of the Sri Lankan health care system.

Sri Lanka recorded its first case of COVID-19 on the 27^th^ January 2020. The case was a Chinese national who came to Sri Lanka as a tourist. The case was successfully treated and discharged [8]. The second case was detected on 11^th^ March 2020, a contact from an European tourist, some six weeks after detection of the first case [9]. As of 4^th^ of April 2020, Sri Lanka has recorded 159 COVID-19 cases, with five deaths [10].

Data from Sri Lanka on COVID-19 shows that it is still within a specific cluster (4^th^ of April 2020) where contact history can be traced; those who contacted the disease from outside the country, their close relatives or those associated with the index case. However, with increasing number of contacts to a single case, the field level difficulties in tracing each and every contact poses a threat of initiating community transmission. The Sri Lankan government initiated measures to prevent the COVID-19 epidemic even before the first case was detected in January 2020. Those measures were gradually strengthened through mandatory thermal screening at the points of entry to the country, followed by self-quarantine of those returning from designated countries from the beginning of March 2020. Mandatory institutionalized quarantine for arrivals from designated countries started on 10^th^ March. This was followed by self-quarantine for all the contacts and all passengers returning from outside the country. Entry points to Sri Lanka for all passengers was closed on 20^th^ of March. All schools, Higher Education institutions, and later, all public and private workplaces except those for essential services were closed. An island wide curfew was imposed on 20^th^ March to implement strict social distancing. This was relaxed intermittently for a few hours to facilitate access to essential services. These measures continue at the time of writing (figure 1), (S1 Appendix). Hence, Sri Lankan preparedness and preventive/ control measures for COVID-19 were unique compared with most other countries.

**Figure 1.**
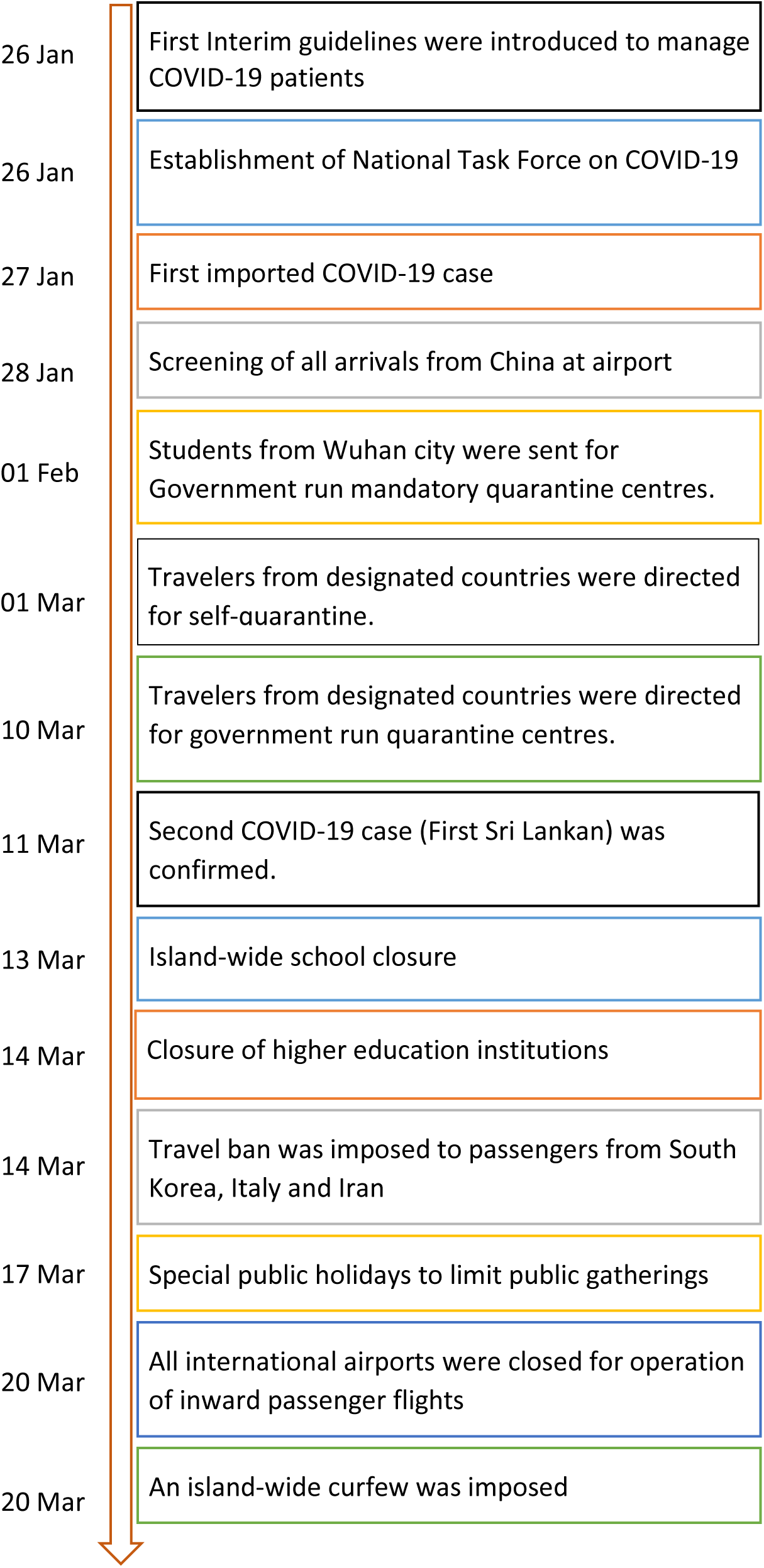
Control measures to mitigate the spread of COVID-19 in Sri Lanka

As stated above, Sri Lanka initiated stringent measures even before a community outbreak of COVID-19 was evident. This makes Sri Lanka a unique place to study the behaviour of this epidemic. Our objective was to develop a mathematical model to predict the epidemic in Sri Lanka incorporating measures taken to implement social distancing, prevent social gatherings, and improve sanitary advice.

## Methods

In this study, a mathematical model was developed and applied to COVID-19 active cases in China, Republic of Korea, and Italy, parameters were also estimated using existing active COVID-19 data from these respective countries where data was available in the public domain [11] The developed model was validated for those three countries. Finally, the model was then applied to the Sri Lankan data from the Epidemiology unit [12], to simulate the epidemic behaviour and obtain projections. The model development, parameter estimation and model validation process are described in detail below.

### Development of Mathematical Model

In the model development process, we assumed that all the clinically tested positive cases for COVID-19 are homogeneous with no impact of age, sex and history of chronic diseases on the disease progression. Conventional compartmental transmission models assume exponential growth for the number of infectious cases during the early stage of a well-mixed epidemic [13,14]. Therefore, the growth pattern of infectious disease outbreaks has been studied using exponential and/or extended versions of exponential models in the absence of control strategies [13–16]. In the present study, we propose a combined hybrid type model which has both exponential and polynomial features with control levels. Generally, exponential models describe either a growth or a decay process depending on the nature of the exponential parameter; polynomial models describe the turning points of a behaviour. The experience gathered from the existing data on COVID-19 shows that pure exponential models may not be able to explain the behaviour of this epidemic. In addition, many externalities, as control measures, also influence the epidemic trend. Hence, we designed a hybrid model by combining both exponential and polynomial models which may be more suited to explain the realistic behaviour of the COVID19 epidemic in the early stages.

Based on the aforementioned theoretical underpinnings, the number of active infected cases (*C(t)*) may be modelled combining quadric and exponential behaviours and is given as follows:

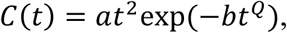

where *C(t)* describes the cumulative number of active cases at time *t*, *a* is a positive parameter, and *b* is either a positive or a negative parameter which depends on trends of the existing data. Parameters *a* and *b* are estimated using the current number of reported active infectious cases. If the current trend shows increasing dominant behaviour we may expect the estimated parameter *b* to be positive. If the current trend shows decreasing dominant behaviour then we may expect *b* to be negative. Therefore, both parameters *a* and *b* are responsible to promote and propagate present trend. However, the situation may change due to the control measures taken by authorities and communities. Those changes can be implemented in the model via parameter *Q*. Parameter *Q* is a free parameter which can be determined based on control strategies and /or community responses.

### Parameter Estimation

Parameters *a* and *b* can be estimated using nonlinear least-squares curve fitting techniques through the active cases curve modelled by *C(t)*.We use the MATLAB in-built function *lsqcurvefit* which enables us to fit parameterized nonlinear functions to data. This function uses the Levenberg-Marquardt algorithm. The parameter Q was chosen by imposing control strategies and/or social distance norms.

## Model Validation

In real time modelling situations, model validation cannot be carried out using traditional techniques as Sri Lanka is still in the initial stages of the epidemic due to having only a small number of cases [16]. However, we studied the behaviour of the COVID-19 epidemic in three countries, namely, China, Republic of Korea, and Italy. These three countries are currently in three different stages of the epidemic; COVID-19 in China is mostly under control, Republic of Korea is likely to be approaching control status, and Italy is still in a full blown community epidemic. We used three data sets from China, Republic Korea, and Italy that are available in the public domain for the purpose of model validation [11] (S2-S4 Appendix).

The objectives of the model validation process are as follows: (1) to determine whether our model captures the epidemic behaviour of the three reference countries, and (2) to determine the effectiveness of control measures, and the duration needed to observe the impact of the control measures on the epidemic curve.

### Model validation using the data from China, Republic of Korea, and Italy

Four simulations were performed using reported active COVID-19 cases from China in four different time periods (Table 1). Similarly, three simulations were performed using active COVID-19 cases from Republic of Korea and Italy in three different time periods (Table 1). Model parameters (*a* and *b*) were estimated in each simulation and predicted using the fitted model. R-square was computed in each trial to evaluate the model fit.

**Table 1.** Time period used for each simulation by country

| Simulation | Time period (in days) |  |  |
| --- | --- | --- | --- |
|  | China | Republic of Korea | Italy |
| 1 | 20-30 Jan (11) | 15-28 Feb (14) | 15-28 Feb (14) |
| 2 | 20 Jan-14 Feb (26) | 15 Feb-14 Mar (29) | 15 Feb-14 Mar (29) |
| 3 | 20 Jan-28 Feb (40) | 15 Feb-28 Mar (43) | 15 Feb-28 Mar (43) |
| 4 | 20 Jan-28 Mar (69) | N/A | N/A |

N/A-Not applicable

Figure 2 presents simulation results for China at four different time periods and the number of active COVID-19 cases reported over time up to 28^th^ March. The red line shows the results of the first simulation. China imposed very strict lockdown conditions on 24^th^ January, and maintained them until the middle of March. The impact of the control strategies cannot be seen in the first 14 days. The green line presents the results of the second simulation. The epidemic still shows an exponential behaviour. However, the gap between the red and green lines shows the impact of the control strategies. The black and blue curves present the results of the third and fourth simulations, respectively. Both curves show a declining behaviour of the COVID-19 epidemic.

**Figure 2.**
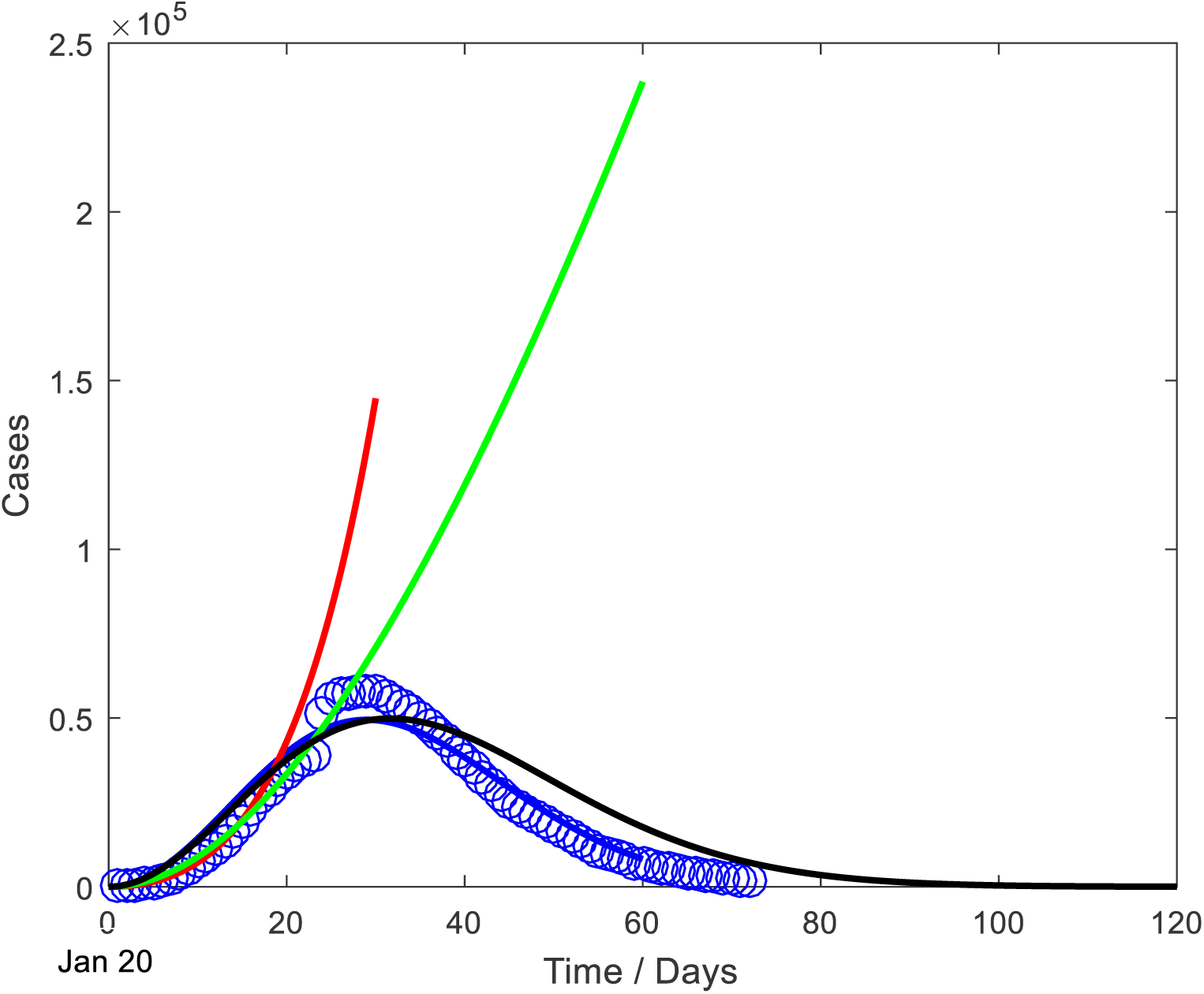
Simulation results and active COVID-19 cases reported in China Red line-Simulation based on 20-30 Jan Green line-Simulation based on 20 Jan-14 Feb Black line-Simulation based on 20 Jan-28 Feb Blue line- Simulation based on 20 Jan-28 Mar Blue spiral line-active COVID-19 cases reported (true data)

Figure 3 presents the simulation results for Republic of Korea at three different time periods and the number of active COVID-19 cases reported over time up to 28^th^ March. The red line corresponds to the results of the first simulation. The curve shows an exponential behaviour. Republic of Korea initiated early and large scale testing and contact tracing to curb the outbreak. However, the impact of the control strategies cannot be seen in the first 14 days. The green and blue lines present the results of the second and third simulations, respectively. Both lines show a declining behaviour of the COVID-19 epidemic.

**Figure 3.**
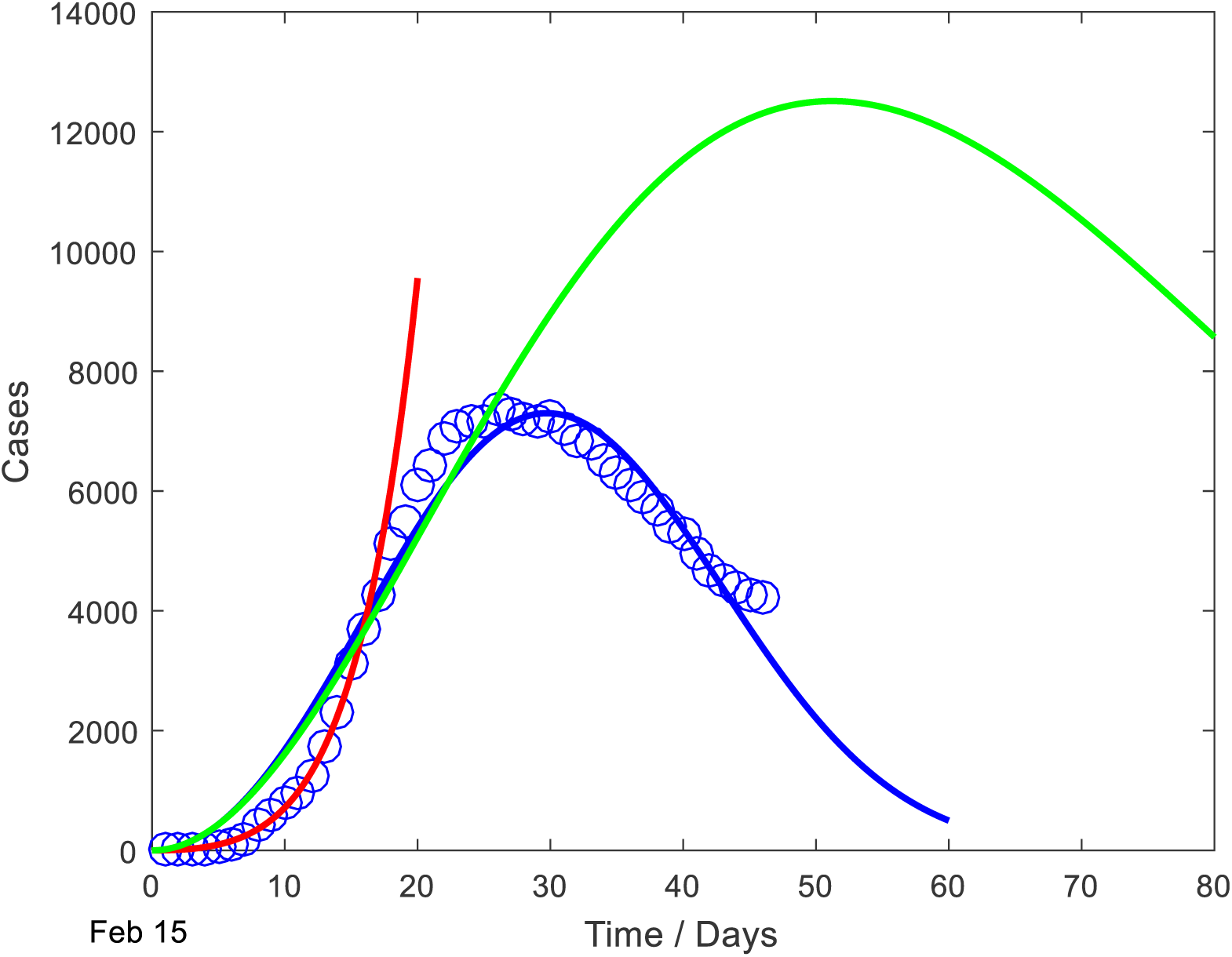
Simulation results and active COVID-19 cases reported in Republic of Korea Red line-Simulation based on 15-28 Feb Green line-Simulation based on 15 Feb-14 Mar Blue line- Simulation based on 15 Feb-28 Mar Blue spiral line-active COVID-19 cases reported (true data)

Figure 4 presents simulation results for Italy at three different time periods and the number of active cases reported over time up to 28^th^ March. The red line corresponds to the results of first simulation. The curve shows an exponential behaviour. Italy imposed several control measures to curtail the outbreak. However, the impact of the control strategies cannot be seen in the first 14 days. Green and blue lines present the results of the second and third simulations, respectively. Both lines show an exponential growth of the epidemic curve.

**Figure 4.**
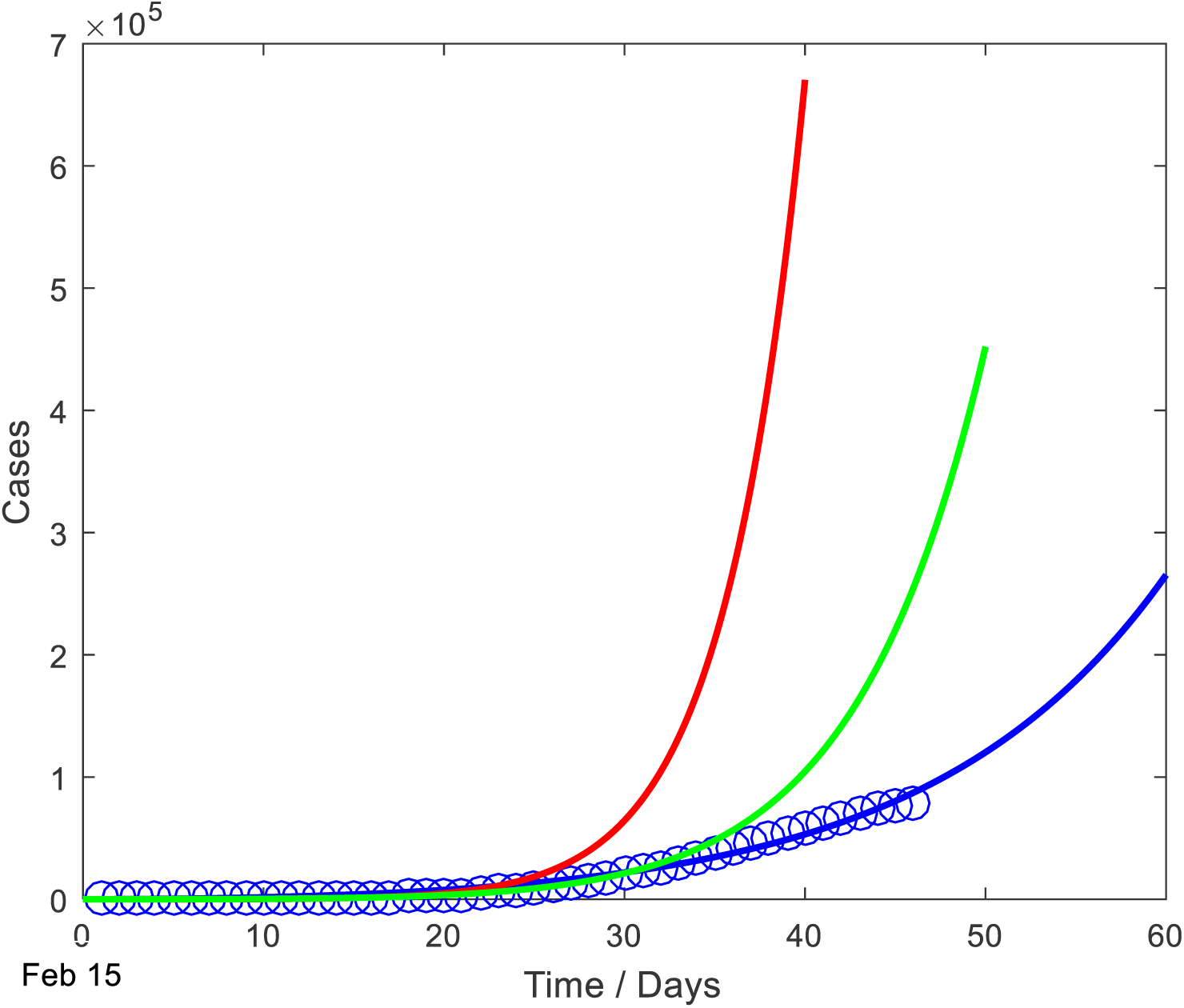
Simulation results and active COVID-19 cases reported in Italy Red line-Simulation based on 15-28 Feb Green line-Simulation based on 15 Feb-14 Mar Blue line- Simulation based on 15 Feb-28 Mar Blue spiral line-active COVID-19 cases reported (true data)

In the graphs of all three countries, the gaps between the curves show the impact of the control strategies- i.e. how effective such controls were and the time duration needed for these to be effective.

The R-square values of all simulated models are higher than 0.91 and are presented in Table 2.

**Table 2.**
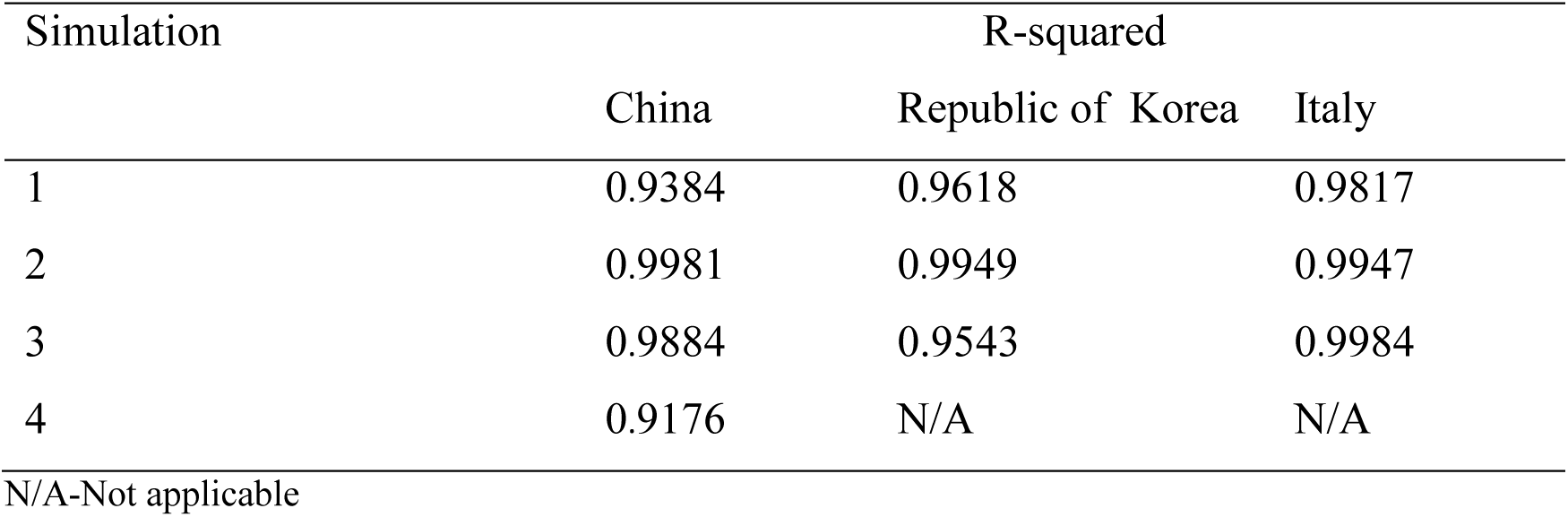
R-square values of simulated models

Finally, the validated model was implemented on the Sri Lankan dataset to simulate the epidemic to obtain projections. The results are described below.

## Results

Simulations were performed using the reported active COVID-19 cases obtained from the Epidemiology unit, Sri Lanka, for the following period: 11–30 March 2020. Figure 5 shows the simulation results and the number of active COVID-19 cases reported in Sri Lanka up to 30th of March 2020. The blue dotted line corresponds to the fitted curve and it explains more than 98% of the observed data (R-squared value =0.9828). According to this projection, Sri Lanka will have a peak of 177 COVID-19 cases by 11^th^ April 2020 (at the end of the second incubation period) if the same trend continues (figure 5-blue dotted line).

**Figure 5.**
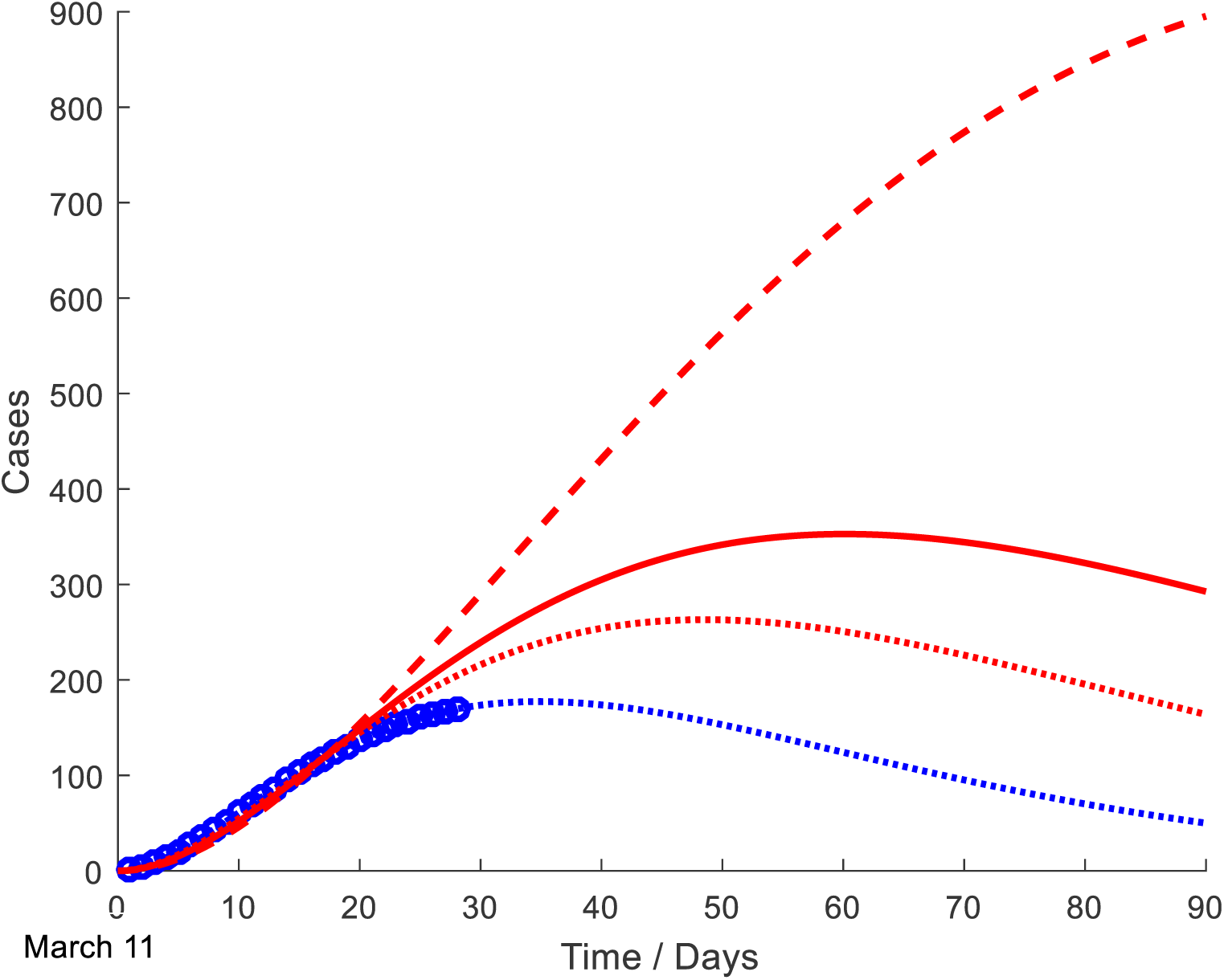
Simulation results for different risk levels and active COVID-19 cases reported in Sri Lanka Red dash line-simulation with 20% risk Red line-simulation with 15% risk Red dotted line- simulation with 10% risk Blue dotted line- simulation for present situation without a risk definition Blue spiral line-active COVID-19 cases reported (true data)

We further defined different risk levels based on varying number of contacts per active infectious case (contact cluster size for each reported case) considering the present control environment (Sri Lanka is currently under curfew [from 20^th^ March] and contacts of each newly identified COVID-19 case are directed to self-quarantine or sent to government run quarantine centres). Contact cluster sizes for this analysis was based on the data received up to the 102^nd^ case reported in Sri Lanka.

A risk of 10% is defined when the number of active contacts of a COVID-19 case is less than 10 along with the number of extended contacts (secondary contacts of each active contact) being 20 or less, and when all the active contacts are kept under quarantine conditions. A risk of 15% is defined as when the number of active contacts are between 10-20 along with number of extended contacts are 10 or less, and when all active contacts are kept under quarantine conditions. A 20% risk is when the number of active contacts is 20 or more along with the number of extended contacts are 20 or less, and when all active contacts are kept under quarantine conditions (Figure 6) [17]. At 10% risk, Sri Lanka will be projected to have a peak of 263 COVID-19 cases by 24^th^ April 2020 (Figure 5-red dotted line) and for 15% risk, the peak will be 353 active cases on 8th May (red line). When the risk level reaches 20% according to the simulation, the peak of the curve will be above 1000 active cases after 90 days.

**Figure 6.**
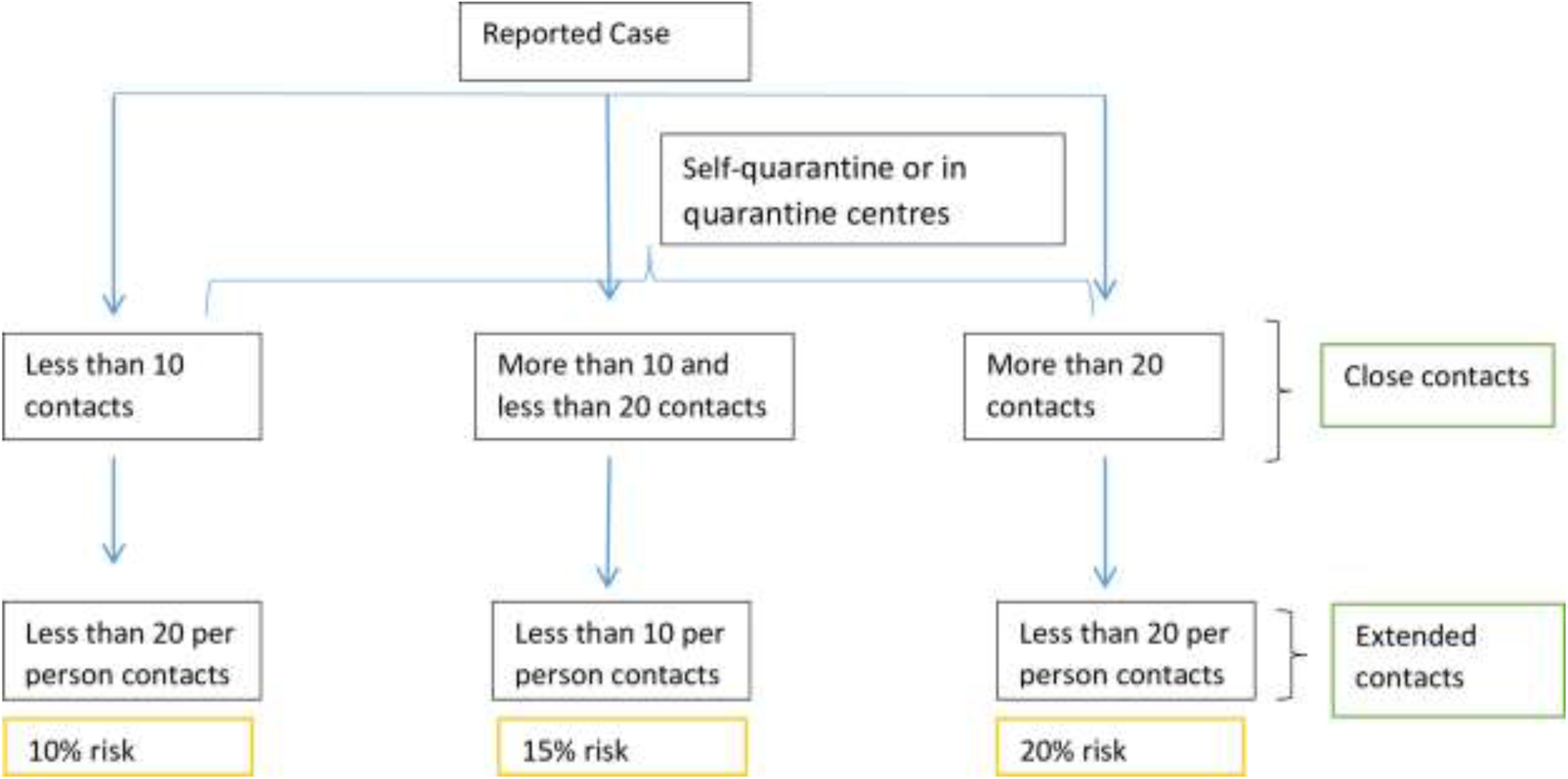
Definition of risk levels

Figure 7 presents the curves shown in Figure 5 separately for each occasion.

**Figure 7.**
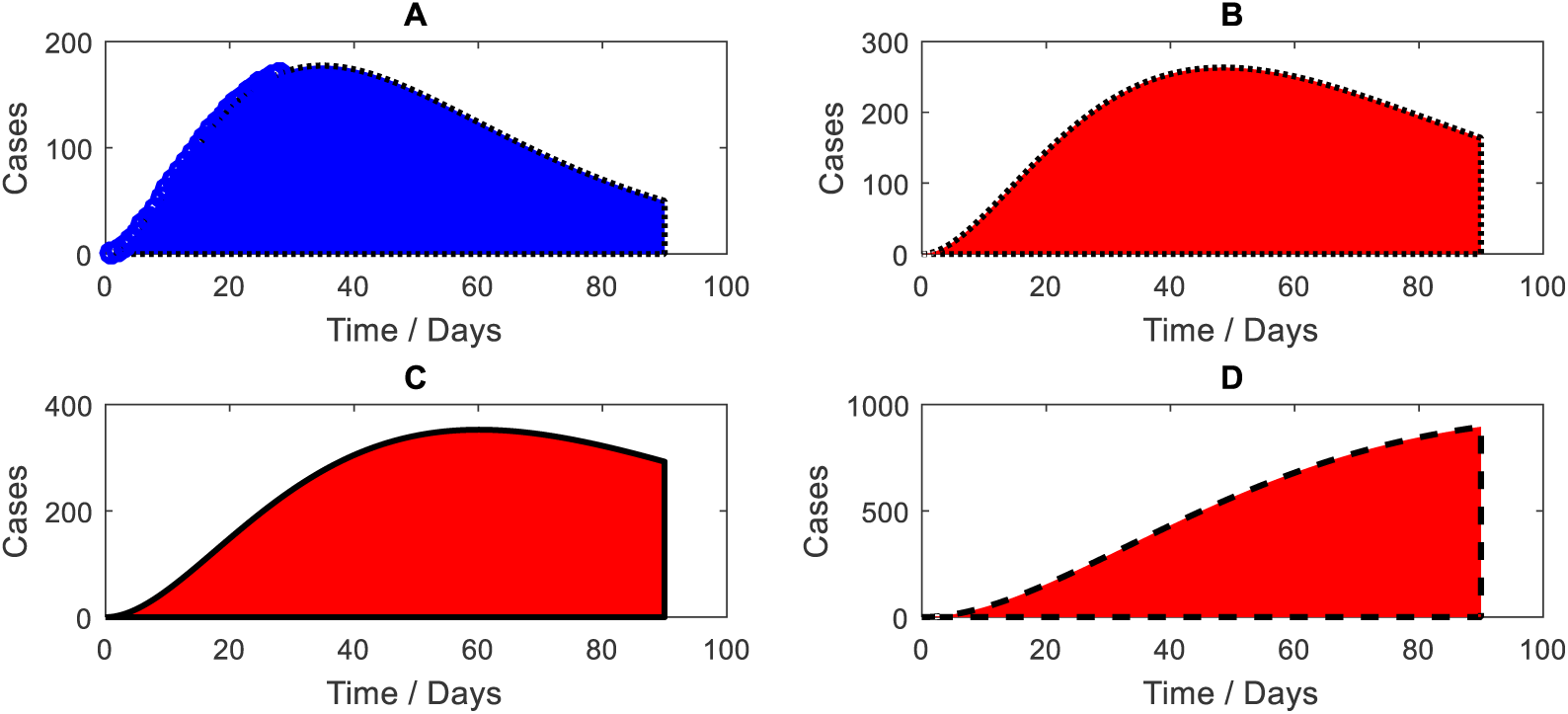
Simulation results for different risk levels and active COVID-19 cases reported in Sri Lanka-expanded A- Simulation for present situation without a risk definition B- Simulation with 10% risk C- Simulation with 15% risk D- Simulation with 20% risk

Figure 8 presents simulations performed according to the degree of different preventive measures targeting social distancing and hygiene. We considered three main factors as control measures, namely keeping social distance (*l_1_*), prevention of social gatherings (*l_2_*) and practising sanitary advisors (*l_3_*). Preventing social gatherings is considered to be a result of imposing curfew where routine activities that enable close contact are totally prohibited. Keeping social distance of one metre is enforced even when the communities are provided with opportunities to obtain essential services to prevent the spread of infection. We scaled down those three factors into percentages of adherence; with ideal status considered as 100% adherence. The combined effect of these three control measures may be modelled using an algebraic product as follows:

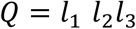

If three conditions are at the maximum control level (i.e. *l_1_ =1, l_2_ =1, l_3_=1)*, keeping social distance of more than 1 metre (100%), prevention of social gatherings (100%), and full adherence to sanitary advice (100%), at maximum control of current measures Q is equal to 1.0. Similarly, if the three measures are at 75% adherence, (0.75*0.75*0.75) then Q is approximately 0.5, and if adherence of all measures is at 50%, (0.5*0.5*0.5), then Q is approximately 0.1. The simulations presented in figure 8 show that if the current measures are fully adhered to (100%), then the number of active cases depicted by the dotted blue line stabilizes (177 COVID-19 cases by 11th April 2020) and then declines. However, deviations from 100% adherence, as depicted by the dotted red (75%) and red (50%) lines, show an increasing trend of active cases that approaches an exponential behaviour.

**Figure 8.**
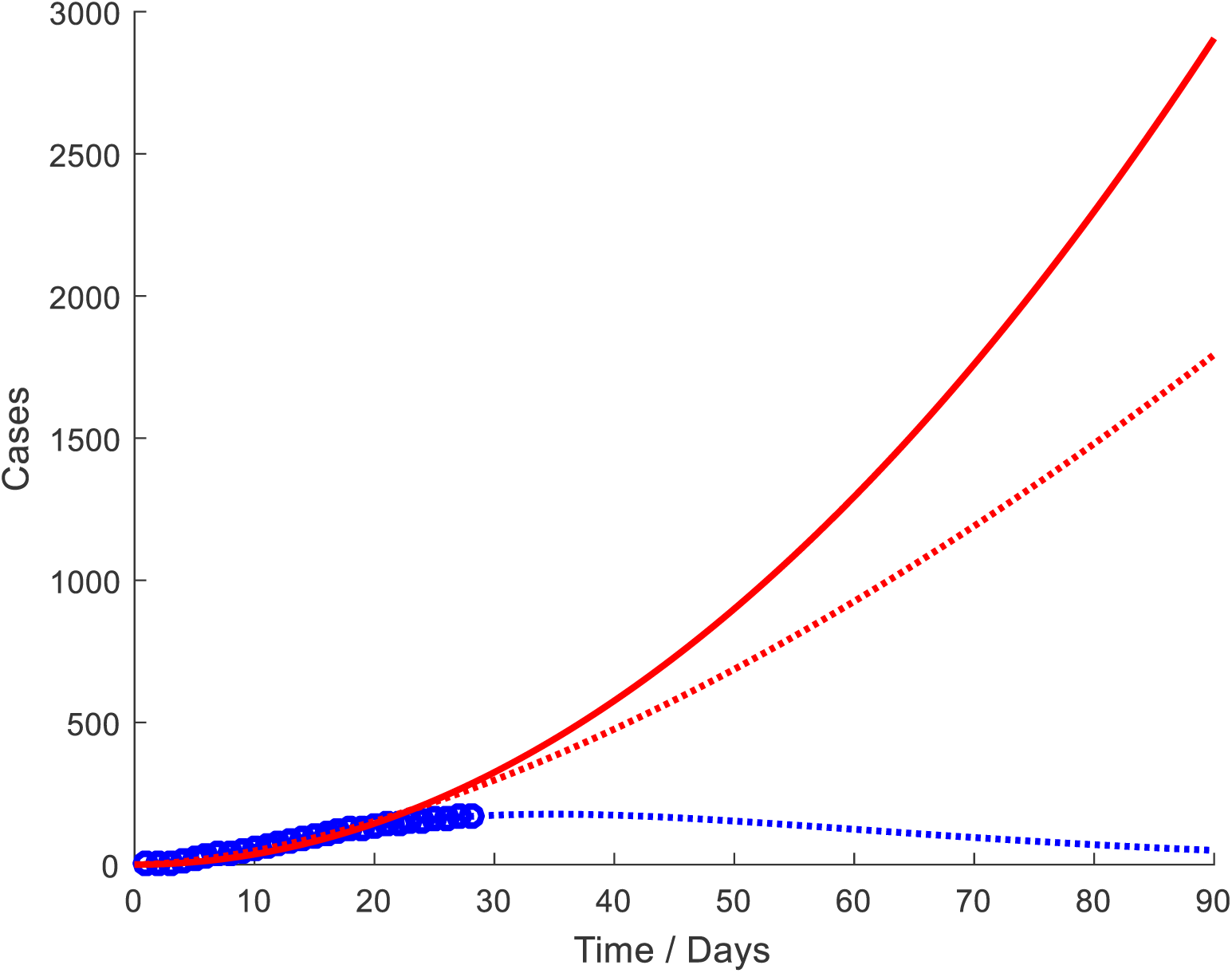
Simulation results for different degrees of control measures and active COVID-19 cases reported in Sri Lanka Red line-Q=0.1 Red dotted line- Q=0.5 Blue dotted line- Q=1.0 Blue spiral line-active COVID-19 cases reported (true data)

## Discussion

A combined hybrid type mathematical model which has both exponential and polynomial features incorporating control levels was developed and validated based on three real time datasets from China, Republic of Korea, and Italy. An excellent model fit was observed for those three reference countries. This model was applied to the limited Sri Lankan data set of COVID19 cases reported from 11th March to 30^th^ March 2020 with a caseload of 122 diagnosed cases. At the time of model development, Sri Lanka still shows a cluster epidemic within the returnees from overseas, and their immediate and extended contacts. Our projections showed a peak of 177 active cases on 11th of April if the current trend continues. However, when additional risk levels of 10%, 15%, and 20% are applied based on the size of the contact clusters, the peak levels of the epidemic shown by the projections stand as 263 active cases by 24th April 2020 (10% risk), 353 active cases on May 8th (15%), and above 1000 active cases after 90 days (20% risk). In addition, our model projects that the direction of the epidemic curve will change depending on relaxation or on the degree of compliance to the current control measures, resulting in an exponential behaviour for the scenarios of lower compliance.

Earlier this year, Sri Lanka imposed gradual control measures to prevent COVID-19 imported cases, even before the first case was detected in late January. The first case did not establish local transmission of the disease and the second case, a contact with a foreigner, was detected six weeks later. Following the detection of a second COVID-19 case and the arrival of more infected returnees from overseas, initiation of local transmission was seen among their immediate contacts establishing a local cluster epidemic. This observation goes along with a recent study which found that a single case introduced to a new location would not necessarily initiate an epidemic, but that several introductions are needed for an outbreak to occur [4]. However, due to the measures already taken, namely self or institutionalized quarantine, those sporadic cases did not spill over to the community, thereby restricting the transmission to within a group of immediate contacts. This allowed Sri Lankan authorities to trace and quarantine the immediate active contacts, and their extended contacts imposing social distancing [17]. Those strict mitigation strategies helped to contain the epidemic within a close community. This is a unique situation that allowed Sri Lanka to observe a cluster epidemic still on 4^th^ of April 2020. Predicting the behaviour of the COVID-19 epidemic in this unique situation incorporating the potential risk of infection spilling over to the community through extended contacts needed a fresh approach. Development of our model through a hybrid approach fulfilled this necessity. Validation of a mathematical model improves the credibility of projections on the future behaviour of the epidemic. Furthermore, it enhances usefulness of predictions to decision makers. We used existing data from three countries to test our model with promising results (R-square above 0.9 for all simulations)[14–16].

The infectivity, thus the basic reproduction number (R_0_) of an epidemic could change with time, and R_0_ will have more reliable estimates with more case data and data on the effects of interventions. In a review examining COVID-19, R_0_ estimates ranged from 1.5 to 6.47 [5]. Data sets in Sri Lanka are limited and so we cannot reliably estimate R_0_ at present, hence our need to rely on published data from other countries. Transmission of COVID-19 to immediate contacts and to their extended contacts will have a bearing on the propagation of the epidemic. In consideration of the available estimates of R_0_ and the contact cluster sizes observed in Sri Lanka, the risk levels were defined as 10%, 15%, and 20% in order to simulate the projections. This allowed us to simulate three additional epidemic curves besides the projections based on the limited existing data. Projections based on different scenarios supports advance planning of the ongoing public health response.

The effects of current public health measures towards the behaviour of the epidemic is important for decision makers. Continuity or discontinuity of such measures warrant evidence [2]. In the absence of many options to mitigate the COVID-19 epidemic, it is vital to examine the available methods to manage and reduce the caseload that are currently in use [3]. Hence, in our simulation, we used three measures already enforced in Sri Lanka; social distancing, prevention of social/community gatherings through curfew, and hygiene practices. Our projections clearly show that the COVID-19 epidemic in Sri Lanka has a slow progression under control conditions. However, based on the three defined risk levels (variation in the levels of contacts being infected), the active caseload shows an increasing trend. Under a scenario of 20% risk, the trajectory of the number of active infected cases curve show stabilizing trend only after 90 days (over 1000 active cases) from the initial case. The results of the projections thus shows the importance of preventive measures to minimize transmission to immediate contacts and thereby reducing the possibility of secondary transmission to extended contacts. Failure to prevent this transmission could lead to a spill over of the infection to the community. When the number of active cases increases, isolation and protective procedures will be less effective to control the epidemic, needing interventions in the curative side [18]. In addition, practical difficulties would also arise in implementing effective quarantine for even the immediate contacts.

Non pharmacological intervention based on sustained physical distancing have a strong potential to reduce the magnitude of the epidemic curve and lead to a small number of total cases [2]. In practice, physical distancing has been implemented through closing of schools and other educational institutions, closure of non-essential work settings and allowing employees to work from home, advocating social distancing through prevention of social gatherings, and imposing curfew or physical lockdown of geographical territories [3,4,18]. Sri Lanka imposed an island-wide curfew on 20^th^ March after nine days of detecting the first active case in our dataset (second case in Sri Lanka) to maximize physical distancing. Although curfew was relaxed for essential services for few hours in between, the trend of the epidemic curve accommodated the effect of those measures during the last ten days of our data set (20-30^th^ of March 2020). Thus, the parameters in our model captured the effects of those control measures in the simulation to provide a reasonable projection. Our projection considered a 100% adherence to those measures to define Q as 1.0. However, realizing that behavioural variations need to be accommodated, particularly on keeping at least one metre distance, the breaching of social gathering prohibitions when curfew is relaxed, and by citizens not fully complying on hygiene practices, separate simulations were done to examine the epidemic behaviour. This is to provide additional time and opportunity for healthcare systems to respond [2].

The projections have an additional utility in deciding when and how to relax control measures, particularly physical distance maintenance, to prevent another surge in the epidemic. The limited literature available at present suggest that a premature and sudden lifting of interventions could lead an early secondary peak and that this can be prevented by a staggered lifting of control measures [2,3]. Our projections on relaxing or noncompliance of control measures clearly confirm this upward surge. Hence, Sri Lanka needs to be extra cautious when making decisions on the existing control measures.

## Strengths and limitations

This is the first known prediction model for the COVID-19 epidemic in Sri Lanka based on local data and case contact data. The model was validated using three large datasets prior to application on a small Sri Lankan data set. The model showed excellent goodness of fit with all three data sets. Additionally, incorporating control measures to the model helped us to examine the contribution of those in mitigating the epidemic.

The dataset in Sri Lanka is still relatively small, hence traditional in built methods of model validation were not possible. Further, incorporation of cluster size of contacts into the model had to be limited to three categories due to the small numbers in the dataset. Our projections based on a homogenous population could be limited to some extent due to the varying behaviour of transmission within different clusters owing to demographic, health seeking, and cultural practices.

### Study implications

Projections of this model are already being used for the mitigation activities of COVID-19 by the Sri Lankan health authorities. These projections have supported decision makers to plan testing facilities, health care provision and human resource management strategies. Projections are regularly updated with new information on active cases and their contacts. The methodological approach used in developing this method can be replicated in other settings with low caseload at the initial stages of the epidemic to project their behaviour, which will be useful for decision makers.

## Conclusions

In conclusion, the hybrid model developed combining exponential and polynomial functions showed promising results to predict the trends of the COVID-19 epidemic. Further, additional simulations incorporating control measures clearly showed that deviations in the levels of compliance could trigger a higher number of active cases and bring upon the exponential features of the epidemic. At this stage our evidence indicates that early relaxation of the control measures that are in place is not advisable.

## Data Availability

Data are available from the authors upon a reasonable request.

## Contributors

SNP and MCW conceptualized the study. SNP and NCG performed mathematical modelling. MCW and DDS provided epidemiological inputs. SNP, MCW, DDS, and NCG interpreted the results and wrote the article.

## Acknowledgements

We would like to thank Dr. Sumudu Karunarathne for helping to locate government publications on COVID-19 and Dr Shaun Scholes, Research Department of Epidemiology and Public Health, University College London for reviewing our manuscript and providing expert comments. We would also like to acknowledge Sri Lanka Medical Association and Epidemiology Unit of the Ministry of Health, Sri Lanka for their support in numerous ways.

